# Non-invasive lung cancer diagnosis and prognosis based on multi-analyte liquid biopsy

**DOI:** 10.1101/2020.08.20.20179044

**Authors:** Kezhong Chen, Jianlong Sun, Heng Zhao, Ruijingfang Jiang, Jianchao Zheng, Zhilong Li, Jiaxi Peng, Haifeng Shen, Kai Zhang, Jin Zhao, Shida Zhu, Yuying Wang, Fan Yang, Jun Wang

**Affiliations:** Department of Thoracic Surgery, Peking University People’s Hospital, Beijing, 100044, China; Envelope Health Biotechnology Co. Ltd., BGI-Shenzhen, Shenzhen, 518083, China; BGI Genomics, BGI-Shenzhen, Shenzhen, 518083, China; Shenzhen Engineering Laboratory for Innovative Molecular Diagnostics, BGI-Shenzhen, Shenzhen, 518120, China

**Author notes:** Corresponding authors: Yuying Wang, Fan Yang, and Jun Wang. Authors contributed equally to this work (Kezhong Chen, Jianlong Sun, Heng Zhao).

## Abstract

Chest LDCT provides an effective approach for lung cancer screening, yet has been found to generate a large number of false positives during practice due to excessive diagnosis of pulmonary lesions of indeterminate clinical significance. In this study, we performed comprehensive genetic and epigenetic profiling of cfDNA from lung cancer patients and individuals bearing benign lung lesions, using ultra-deep targeted sequencing and targeted bisulfite sequencing. We found that cfDNA mutation profile alone has relatively limited power in distinguishing malignant from benign plasma, while cfDNA methylation profiling showed a better performance for classification of the two groups and combination of genetic and epigenetic features of cfDNA along with serum protein marker further improved the classification accuracy. We also identified novel methylation-based prognostic markers and showed that an integrated model that combined cfDNA mutational status and methylation-based prognostic markers improved prediction for lung cancer survival. Our results highlight the potential of the multi-analyte assay for non-invasive lung cancer diagnosis and prognosis.

## Introduction

Lung cancer (LC), with the highest incidence and mortality rates among cancers worldwide, is the leading cause of death in many countries including China [1]. The stage at which lung cancer is diagnosed has a significant impact on the prognosis of this disease. A study showed that the 5-year overall survival rate was 57.4% for localized lung and bronchus cancers and merely 5.2% for distant ones [2]. However, timely detection of lung cancer remains difficult since patients are often asymptomatic at an early stage of the disease.

Low-dose computed tomography (LDCT), as a replacement of chest radiography, is the most extensively recommended lung cancer screening method currently [3]. Its effectiveness has been proved by the National Lung Screening Trial (NLST) which demonstrated a relative reduction of 20.0% in lung cancer mortality with this approach [4]. However, LDCT as a screening method poses radiation risk. The cumulative radiation exposure of a participant following the current lung cancer screening protocols over 30 years could reach 420 mSv, which exceed those among nuclear power workers as well as atomic bomb survivors [5]. Additionally, the false-positive rate of LDCT can be up to 50%, while the positive predictive value could be as low as 2.4% [6]. This is due to the difficulty in distinguishing between malignant and benign lung nodules by CT scans [7]. The resultant overdiagnosis and overtreatment could potentially lead to adverse medical events [8]. Therefore, new screening technologies for overcoming these drawbacks are required.

Derived from tumor cells, circulating tumor DNA (ctDNA) in plasma of cancer patients provides valuable information about cancer and also holds great promise for non-invasive early cancer detection [9–13]. However, since ctDNA is diluted by circulating cell-free DNA (cfDNA) of noncancerous origins, its detection poses significant challenges especially during early stages of cancer when the tumor mass is small [14,15]. Notably, ctDNA contains both genetic and epigenetic information that may derive from the tumor, including but are not limited to mutation spectrum, copy number variation (CNV), changes in genomic methylation level, and fragmentation patterns [13,16–18]. Therefore, it is an attractive hypothesis that simultaneous analysis of multiple features may improve ctDNA detection. Nevertheless, previous studies on early cancer detection have mostly focused on a single feature of the ctDNA, such as cancer driver gene mutations or alterations in the methylome [14,19–21].

In this study, we have developed a set of experimental and computational tools to measure both genetic and epigenetic signals from plasma cfDNA of lung cancer (LC) patients as well as patients bearing benign lung lesions (BLN) using high-throughput sequencing, aiming to explore the potential utility of blood-based biomarkers for lung cancer diagnosis and for prediction tumor recurrence risk.

## Methods

### Patients enrolled and samples collected in this study

Between December 2013 and December 2018, 128 LC and 94 BLN patients were enrolled in this study at the Peking University People’s Hospital, Beijing, China, with the informed consent form signed by every participant. This study was approved by the Ethics Committee of Peking University People’s Hospital (No.2017PHB106–01). The histopathological classification was based on the 2015 World Health Organization classification [22]. 4–8 mL blood was collected from the participants before surgery into 10 mL K2EDTA tubes (BD, 366643) and stored at room temperature. Plasma separation was performed within 4 hours after collection by centrifugation at 1,600×g for 10 minutes and then at 16,000×g for another 10 minutes at room temperature. Separated plasma was stored at –80 °C until DNA extraction. 25 pairs of lung cancer tissues and adjacent normal tissues were collected during surgery at stored at –80 °C.

### DNA Extraction and Quality Control

Plasma cfDNA extraction was conducted by MagPure Circulating DNA Maxi Kit (Magen, 12917PC-100) following the manufacturer’s instructions with some modifications. The concentration of cfDNA was measured using the Qubit™ dsDNA HS Assay Kit (Thermo Fisher Scientific, Q32854). The quality of cfDNA was analyzed by Agilent High Sensitivity DNA Kit (Agilent Technologies, 5067–4626) and Agilent 2100 Bioanalyzer (Agilent Technologies). cfDNA samples with excessive high molecular weight nucleic acids were considered as contaminated by white blood cell genomic DNA (WBC gDNA) and were excluded from further analysis. gDNA was extracted from WBC, lung cancer tissues, and normal tissue adjacent to the tumor (NAT) using MagPure Buffy Coat DNA Midi KF Kit (Magen, D3537–02) per manufacturer’s instruction, and DNA concentration was measured by Qubit™ dsDNA HS Assay Kit.

### Capture panel design for targeted ultra-deep Next Generation Sequencing (NGS)

We used a 139-gene pan-cancer panel for targeted ultra-deep sequencing. Targeted genes and exons were selected based on mutation frequency in the The Cancer Genome Atlas (TCGA) database [23] and the COSMIC database of somatic mutations in cancer[24], prioritizing cancer driver genes [25]), and exons with TCGA or COSMIC hotspot mutations.

### Library preparation for targeted ultra-deep NGS

To reduce noises that may derive from PCR and/or sequencing errors, we used a duplex unique molecular identifier (UMI) strategy in library preparation, adapted from a previous study [26]. Briefly, cfDNA was end-repaired and ligated to sequencing adapters, and index PCR was performed followed by purification by Agencourt AMPure XP beads (Beckman Coulter, A63882). WBC gDNA was processed in the same way except for it was fragmented by sonication before library preparation.

Target capture reactions were performed using xGen^®^ Lockdown^®^ Reagents (IDT technologies) per manufacturer’s instruction. Captured Libraries were amplified in a 50 μL PCR mix composed of 25 μL 2× KAPA HiFi Hot Start Ready Mix, 5 μL PCR primer pair (10 μM) and 20 μL beads suspensions with the following cycling conditions: 45s at 98°C, followed by 13 cycles of 98°C for 15 s, 60°C for 30 s, and 72°C for 30 s; final extension was performed at 72°C for 1min. Libraries were purified by Agencourt AMPure XP beads, quantified by Qubit^TM^ dsDNA HS Assay Kit, and sequenced on MGISEQ-2000 (MGI Tech) using 2×100 paired-end sequencing.

### Library preparation for targeted bisulfite sequencing

To improve the quality of cfDNA whole-genome bisulfite sequencing (WGBS) libraries, we adopted a single-stranded DNA (ssDNA) library preparation strategy. Briefly, bisulfite conversion was performed on input DNA using EZ DNA Methylation-Gold^TM^ Kit (Zymo Research, D5006) per manufacturer’s instructions. Next, bisulfite-converted ssDNA was ligated to sequencing adaptors as described previously [27]. gDNA extracted from lung cancer or normal tissues was fragmented by sonication before library preparation.

Targeted capture reactions of the WGBS libraries were performed using SeqCap Epi CpGiant Probes (Roche) following the manufacturer’s instruction. Captured libraries were amplified and sequenced on MGISEQ-2000 using 2×100 paired-end sequencing.

### Variant analysis

Targeted sequencing data from cfDNA libraries were processed as follows: UMI sequences were trimmed from fastq data using in-house scripts and were adapter trimmed and quality trimmed using SOAPnuke-2.0.3 [28]. Reads were aligned against the human reference genome (hg19) using BWA-MEM (version 0.7.17) [29]. Candidate mutations were identified from the aligned reads using a two-step procedure: Firstly, hotspot mutations (defined as point mutations, small insertions and deletions represented in COSMIC database (https://cancer.sanger.ac.uk/cosmic, version 85) with > = 20 cancer cases) were identified using the in-house script and filtered using an allele fraction cutoff of 0.05% (except for indels, which were not filtered). Secondly, non-hotspot mutations were identified using freebayes (version 1.1.0) [30] and filtered using an allele fraction cutoff of 0.05%. These two sets of variants were combined and filtered for potential germline variants (with allele fraction > = 25%) [14]. Variants were further filtered for germline mutations using a custom germline database derived from the ExAC germline variants data [31] and 1000 Genome data [32], as well as a custom false-positive database. Remaining variants were then annotated using VEP (version 95.2–0) [33]. For cfDNA samples, variants were further filtered using the following set of criteria: variants were first filtered to exclude intronic and silent mutations. For the remaining hotspot variants, only those with at least 3 supporting UMI families and at least one supporting duplex UMI family were retained (except for indels). For the remaining non-hotspot variants, only ones with at least 8 supporting UMI families and at least one supporting duplex UMI family, or ones with at least 6 supporting UMI families and at least two duplex UMI families, were retained. Non-hotspot mutations with a SIFT prediction of “tolerated” and a PolyPhen prediction of “benign” were excluded. Finally, within the remaining non-hotspot variants, only those with a SIFT score < = 0.02 and a PolyPhen score > = 0.95, or a PolyPhen score of 1, or a SIFT score of 0, were retained. For WBC samples, no further filtering was applied. To derive the final set of variants for plasma sample, cfDNA variants were filtered with variants identified from the matched WBC sample.

### Mutation scoring system

Variants were classified and weighted according to the following arbitrarily defined tiered scoring system: COSMIC hotspots with more than 500 cancer cases were given a score of 8; TCGA hotspot variants [34] or COSMIC hotspots with more than 100 cancer cases and not in the former class were given a score of 4; COSMIC hotspots with more than 20 cancer cases and not in the former class were given a score of 2; the rest of variants were given a score of 1.

### Methylation data analysis

Targeted bisulfite sequencing data were processed as follows. First, low-quality reads and 3’ sequencing adapters were trimmed by fastp (version 0.19.7) [35]. Then, pair-end reads were aligned to the hg19 reference genome using BitMapperBS (version 1.0.0.8) [36]. Only reads mapped in proper pair to a unique genomic position and spanning an insert size between 30 bp and 500 bp were retained. Next, duplicates were marked with sambamba (v0.6.8) [37]. Finally, methylation rates were calculated as #C/(#C+#T) for individual CpG sites with at least 4x coverage using MethylDackel (https://github.com/dpryan79/MethylDackel, version 0.3.0).

### Identification of differentially methylated regions (DMRs)

A Bayesian hierarchical model was used to detect the differential methylated loci between 25 lung cancer tissues and 25 matched normal tissues (p< 0.001 and delta>0.2) [38]. To account for the spatial correlation of methylation ratio, smoothing was applied to combine the information from proximal CpG sites to identify differentially methylated regions (DMRs). DMRs were defined as the regions satisfying the following criteria: ≥ 50bp, containing ≥3 CpG sites within the region, and ≥80% CpG sites with significant p-values. Only hypermethylated DMRs were used in the subsequent analysis.

### Predictive model construction

Regional methylation ratio was calculated per DMR for each cfDNA sample sequenced by targeted bisulfite sequencing and processed as features by dividing the sum of methylated cytosine by the sum of depth in the DMR. Ten-fold cross-validation was performed to validate random forest models for classifying plasma cfDNA of lung cancer patients from that of patients bearing benign lung nodules using the python package scikit-learn [39].

Feature selections were performed on the training data only, using a feature importance cutoff of 0.008. Random forest models were fitted using the selected DMRs with the parameters: number of trees = 60, depth = 5. The fitted models were then applied in the validation set from which the sensitivity, specificity, and area under the curve (AUC) were calculated. Multi-omics prediction models were trained and validated similarly, except that feature selections were applied to the DMR features only.

### Identification and validation of prognostic markers

To identify methylation-based prognostic markers, samples were randomly divided into a training set and testing set using a 60/40 split. We applied the following procedure to select the potential methylation-related prognostic factors and to fit prognosis model in the training set: we first removed DMRs with a standard deviation< 0.03 from the identified lung cancer DMRs as mentioned above since less variant features provided limited information; we then used the selected DMRs to fit a LASSO Cox proportional hazard model on OS. Through 10-fold cross-validation, we chose the tuning parameter λ when the partial likelihood deviance reached the lowest, from which DMRs were further filtered and the coefficients of the each DMR were obtained. We calculated the methylation-based prognostic score (MPS) for each individual as the sum of the products of the DMR methylation level and its coefficient and combined the mutation score (wSUMAF) with the MPS as the multi-omics score. We then assessed the association of lung cancer prognosis with mutation score and multi-omics score separately in the training set and testing set. Kaplan–Meier curves were plotted for each analysis. Finally, two separate multivariate Cox proportional hazard models were built on wSUMAF only and both wSUMAF and MPS with adjustment of age, stage, histological type and smoking status in the testing set. To avoid information loss through categorization, both wSUMAF and MPS were analyzed as continuous variables in the multivariate Cox regression. To compare the performance of two models, the incident cases / dynamic controls ROC curve was plotted [40]. The R package of glmnet, survival, survminer (https://CRAN.R-project.org/package=survminer), risksetROC were used. The analyses procedure is summarized in the Supplementary Figure 19.

## Results

### Study design and patients

In this study, we performed a comprehensive analysis of sequence alterations and methylation pattern of plasma cfDNA as well as levels of serum protein markers from lung cancer patients and patients bearing benign lung nodules, in order to explore the possibility of using these features to non-invasively distinguish between malignant and benign lung nodules (Figure 1A). Blood samples were collected from 128 lung cancer (LC) and 94 benign lung nodule (BLN) patients (Table 1). As expected, LC patients had significantly higher mean plasma cfDNA level (20.53 ± 1.04ng/ml) than BLN patients (13.78 ± 1.14 ng/ml, p = 2.14E-05, student’s t-test) (Supplementary Figure 1), in accordance with the previous report [41].

**Table 1:** Clinicopathological characteristics of the patients enrolled in this study. LUAD: lung adenocarcinoma. LUSC: lung squamous cell carcinoma. LCC: large cell carcinoma. SCLC: small cell lung carcinoma.

|  |  | LC (N=128) |  | BLN (N=94) |  |
| --- | --- | --- | --- | --- | --- |
|  |  | Number | Percentage | Number | Percentage |
| Gender | Female | 53 | 41% | 48 | 51% |
|  | Male | 75 | 59% | 46 | 49% |
| Age | Median±SD (Range) | 63.00±11.58 (30-86) |  | 55.00±10.49 (18-79) |  |
| Histology | LUAD | 97 | 76% |  |  |
|  | LUSC | 23 | 18% |  |  |
|  | LCC | 3 | 2% |  |  |
|  | SCLC | 5 | 4% |  |  |
| Stage | 0 | 2 | 2% |  |  |
|  | IA | 54 | 42% |  |  |
|  | IB | 29 | 23% |  |  |
|  | II | 17 | 13% |  |  |
|  | III | 19 | 15% |  |  |
|  | IV | 7 | 5% |  |  |
| Smoking History | Smokers | 20 | 16% | 23 | 24% |
|  | Non-smokers | 33 | 26% | 70 | 74% |
|  | Unknown | 75 | 59% | 1 | 1% |

**Figure 1:**
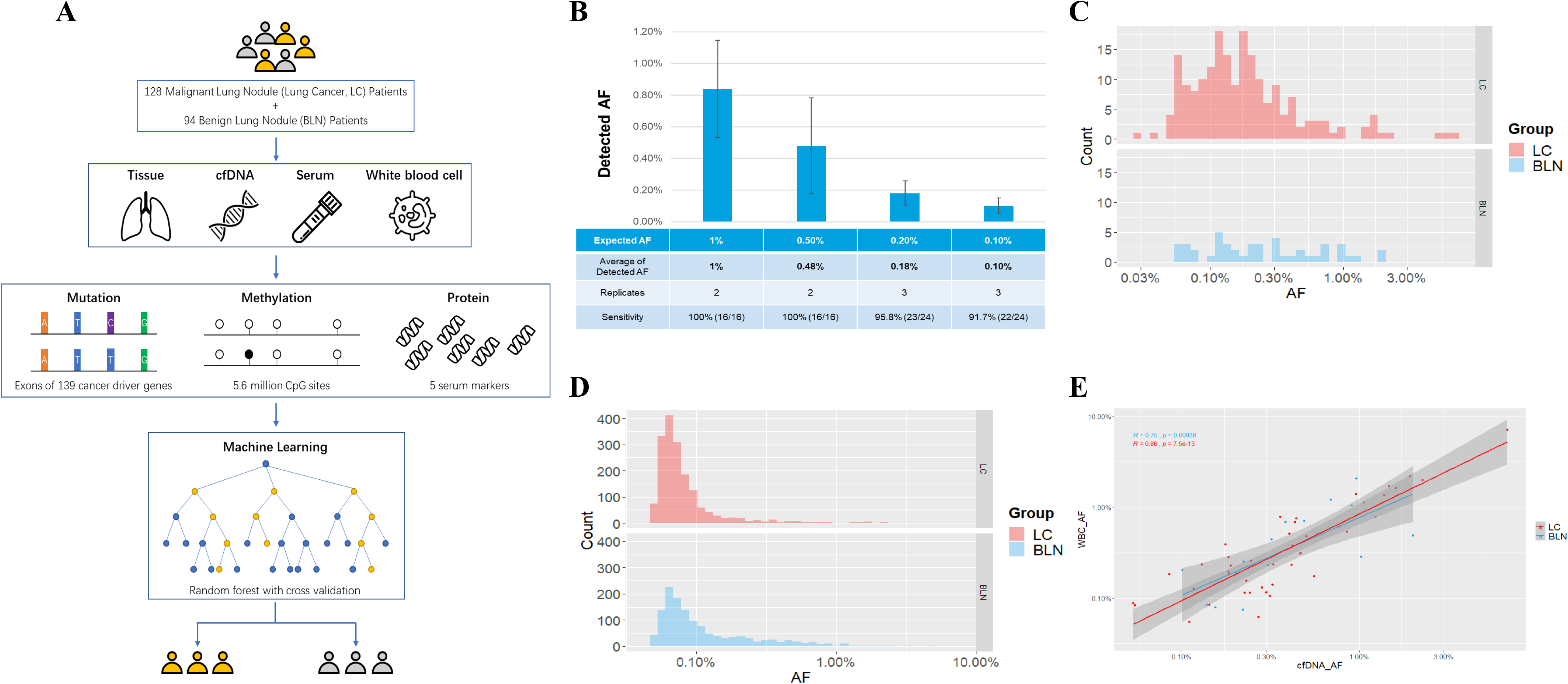
Study design and variants detected by targeted ultra-deep sequencing in cfDNA and WBC gDNA. (A) Schematic view of the study design. See Methods for additional details. (B) Spike-in experiments using Multiplex I cfDNA Reference Standard Set which contains 8 SNVs to test the LOD of our targeted ultra-deep sequencing method. The sensitivity was calculated as the number of detected SNVs divided by the number of total spiked-in SNVs in all the replicates for each condition. (C) Allele fractions (x-axis, log scale) of mutations detected in plasma cfDNA of BLN patients (blue) and LC patients (red). (D) AF (x-axis, log scale) distribution of WBC gDNA variants from BLN and LC patients. E) Pearson correlation of AF in cfDNA (x-axis, log scale) and AF in WBC gDNA (y-axis, log scale). Each point represents one variant detected in matched cfDNA and WBC gDNA samples from the same patient.

### Targeted ultra-deep NGS detected distinct mutational spectrum of plasma cfDNA and WBC gDNA

To profile sequence alterations carried by cfDNA, we performed targeted ultra-deep NGS on plasma cfDNA extracted from 111 LC patients and 78 BLN patients (Supplementary Table 1) using a panel covering exons of 139 cancer driver genes selected based on TCGA and COSMIC databases (Supplementary Table 2, see Methods for panel design). We achieved an average raw target sequencing depth over 50,000× and an average deduped sequencing depth over 5,000× (Supplementary Figure 2). We designed a set of stringent thresholds to identify the most reliable variants, based on the number of supporting UMI families and duplex UMI families, the allele fractions, and function predictions (see Methods for details). Potential germline variants were also removed before downstream analysis. To test the limit of detection (LOD) and evaluate the accuracy of our method, we first performed spike-in experiments using a reference standard containing 8 single-nucleotide variants (SNVs) and cfDNA from two healthy individuals by the method previously reported [42]. Results indicated that our targeted ultra-deep NGS method could efficiently detect mutations with variant allele frequencies (VAFs) of 0.1% and 0.25%, with a sensitivity of 91.7% (22/24) and 95.5% (21/22) respectively (Figure 1B and Supplementary Figure 3). The VAFs of identified sequence alterations using this method ranged from 0.03% to 6.82% with a median of 0.16% for LC patients and from 0.05% to 2.00% with a median of 0.22% for BLN patients (Figure 1C). In total, 193 and 46 mutations were detected in 75 (out of 111, 68%) LC patients and 33 (out of 78, 42%) BLN plasma cfDNA, respectively (Supplementary Figure 4 and 5). As expected, cfDNA of LC patients appears to harbor more sequence alterations than that of BLN patients (Supplementary Figure 6).

**Figure 2:**
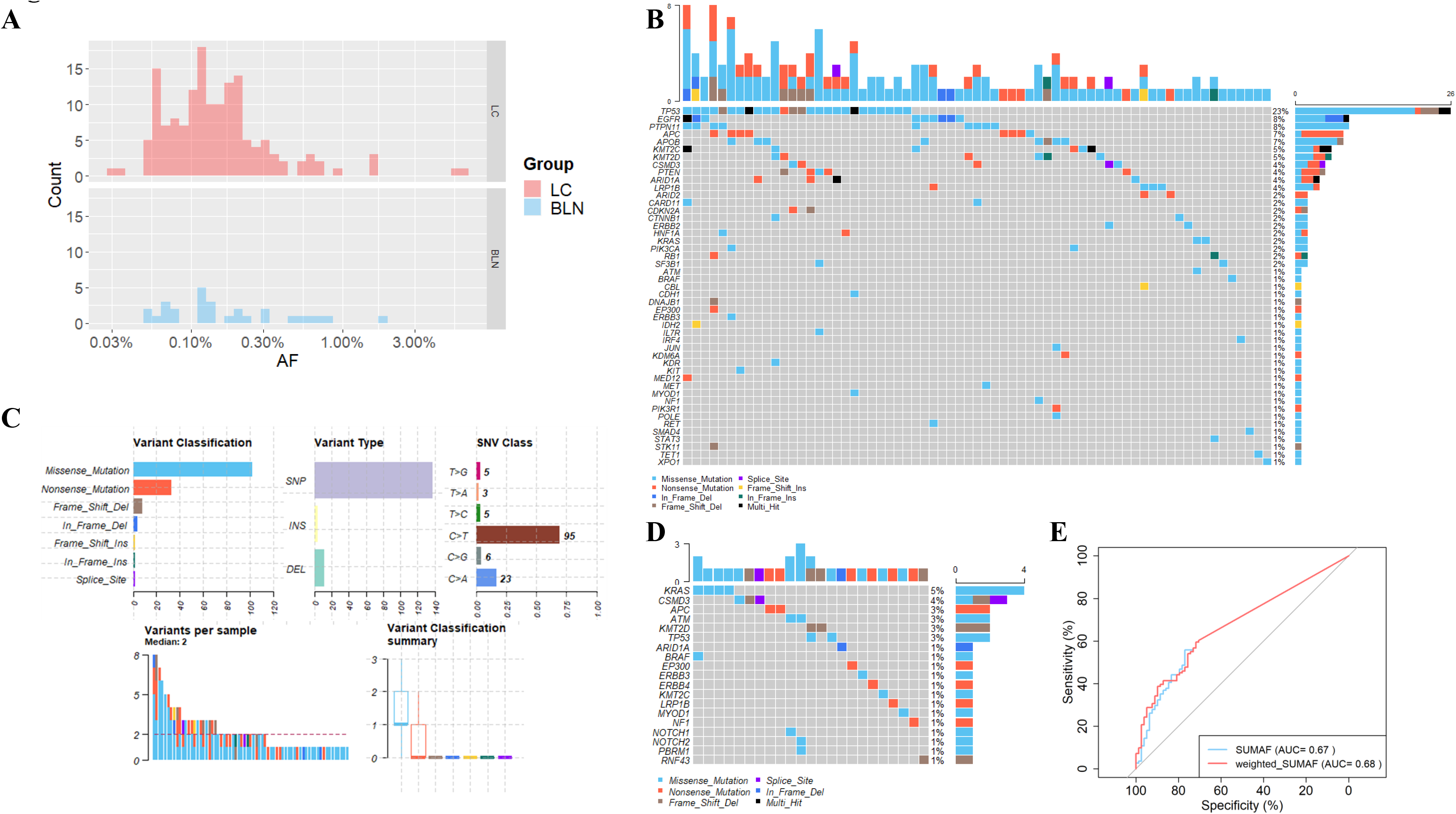
Predictive model based on variants detected in plasma cfDNA after filtering with matched WBC sample for shared variants. (A) AFs (x-axis, log scale) of cfDNA variants of LC patients (red) and BLN patients (blue). (B) Oncoplot showing the 153 mutations detected in 67 out of 111 (60.36%) LC samples. 45 LC samples without any mutation detected were not shown. Each column represents a sample and each row a different gene. The upper barplot represents the frequency of mutations for each sample, and the right barplot represents the frequency of mutations for each gene. Samples are ordered by the most mutated genes. (C) Summary plot of the 153 mutations detected in LC samples. Upper panel from left to right: Variant classification, Variant Type, and SNV Class. Lower panel from left to right: Variants per sample and Variant classification summary. (D) Oncoplot of the 28 mutations detected in 23 out of 78 (29.49%) BLN samples. 55 BLN samples without any mutation detected were not shown. (E) Predictive models to distinguish LC from BLN based on mutations detected. SUMAF (green): sum of AF model. Weighted_SUMAF (red): sum of weighted AF model (see Methods). The AUC of SUMAF model is 0.67 with 55.9% sensitivity and 76.9% specificity. The AUC of weighted_SUMAF model is 0.68 with 59.5% sensitivity and 71.8% specificity.

**Figure 3:**
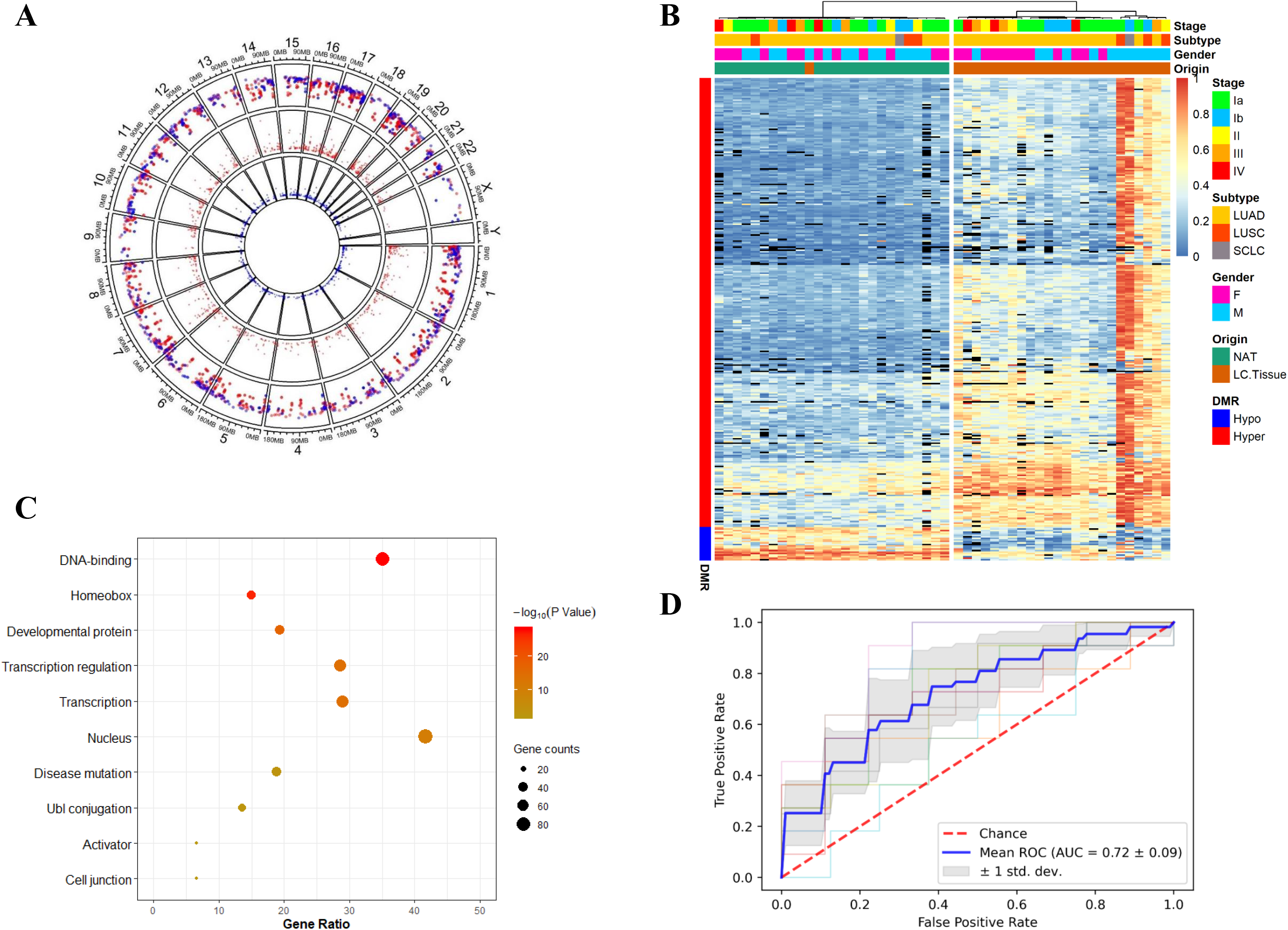
Diagnostic model for Distinguishing LC from BLN plasma by analyzing cfDNA methylation levels. (A) Differentially methylated regions (DMRs) discovered by WGBS of LC tumor and normal tissue adjacent to the tumour (NAT). Red points: Hypermethylated DMRs in LC tissues. Blue points: hypomethylated DMRs in LC tissues. From outer to inner circle, the first circle is overview of DMRs, the second circle is the area statistics of hypermethylation regions (methy.diff>0.2), and the third circle is the area statistics of hypomethylation regions (methy.diff<-0.2). (B) Heatmap of the DMRs with hierarchical clustering. Block color represents the methylation β value and black represents N.A. (C) Functional annotation of the genes associated with the 293 hypermethylated DMRs by gene ontology (GO) terms using DAVID. (D) Predictive models to distinguish LC from BLN based on cfDNA methylation level with selected features (feature importance ≥ 0.008). A total of 76 DMRs were retained in the final model. The AUC is 0.72 with 80.5% sensitivity and 57.5% specificity.

**Figure 4:**
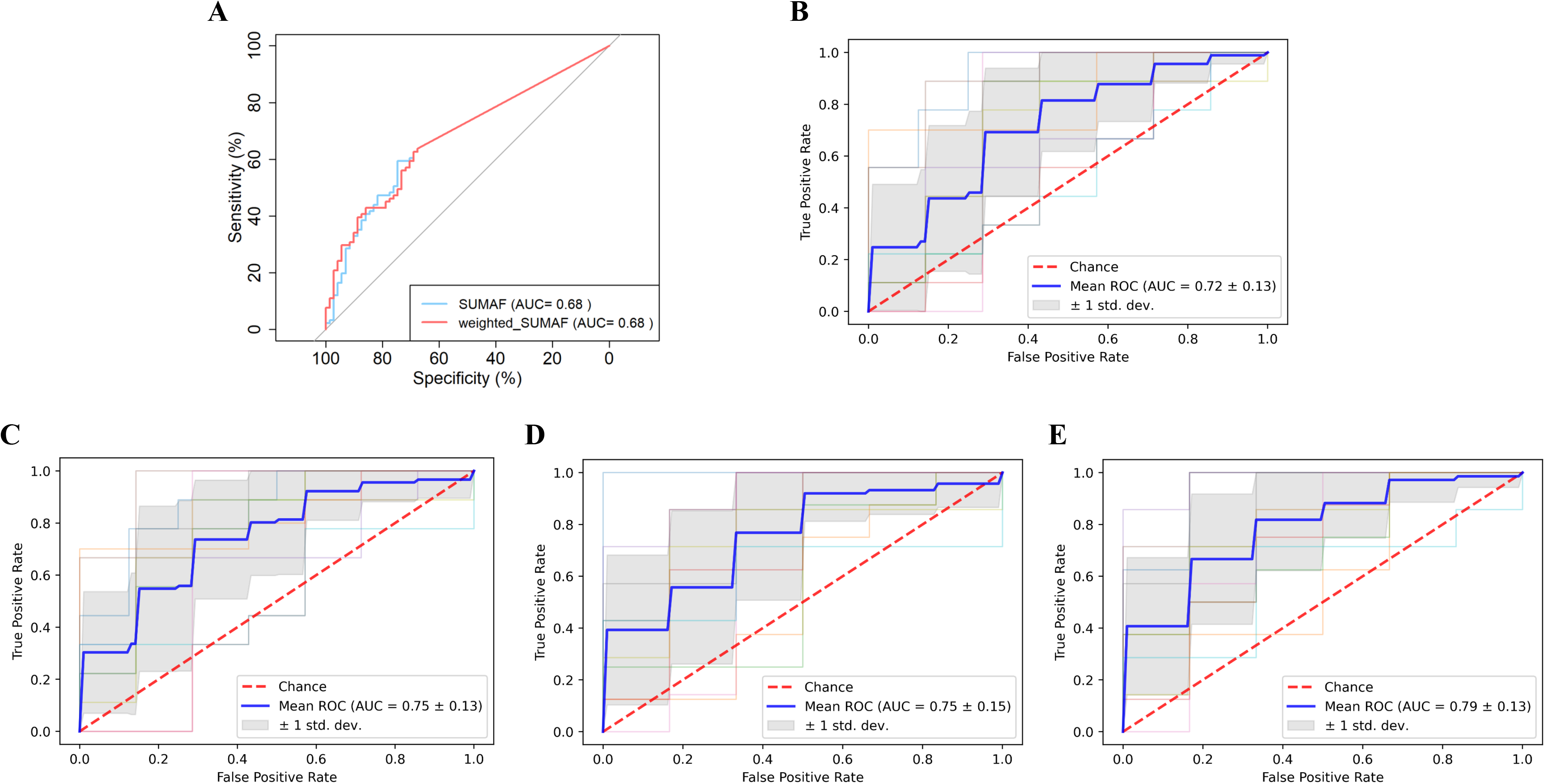
Multiomics predictive models to distinguish LC from BLN plasma. Bi-omics samples: cfDNA samples from 91 LC and 71 BLN patients with complete mutation and methylation data. Tri-omics samples: cfDNA samples from 74 LC and 60 BLN patients with complete measurement of mutation score, methylation levels of selected DMR and serum CEA levels. (A) Classification models built based on mutation status alone in bi-omics samples; (B) Models built based on selected DMRs alone in bi-omics samples; (C) Models built based on mutation score and selected DMR in bi-omics samples; (D) Models built based on mutation score and selected DMR in tri-omics samples; (E) Models based on combined mutation score, selected DMR, and serum CEA levels in tri-omics samples.

Recent studies suggest that some of the variants found in cfDNA may derive from the process of clonal hematopoiesis and confound the analysis [43]. To address this, genomic DNA (gDNA) of white blood cell (WBC) from cfDNA mutation-positive participants were also sequenced with ultra-deep targeted sequencing (see Methods). Non-synonymous variants were detected in WBC of 73 (out of 75, 97%) LC patients and 33 (out of 33, 100%) BLN patients respectively (Supplementary Figure 7 and 8). The AFs of variants observed in WBC samples were mostly less than 1%, ranging from 0.04% to 7.10% (Figure 1D). Within these WBC-shared cfDNA variants, the most frequently mutated genes included *TP53, CBL, APOB* and *CSMD3* for LC plasma, and *CBL, CSMD3*, and *STAT3* for BLN plasma (Supplementary Figure 9). Of these, *TP53* and *CBL* are regarded as canonical CH genes (other canonical CH genes such as *DNMT3A, TET2* and *ASXL1* were not included our targeted panel) [43]. Moreover, AFs of variants shared by plasma cfDNA and matched WBC samples are highly correlated (Figure 1E), suggesting that these mutations indeed originated from WBC and should be removed for downstream analysis. Notably, a number of these mutations were hotspot mutations of cancer driver genes (defined as variants with > = 20 reported cases in the COSMIC database (Supplementary Figure 10). The percentages of cfDNA variants matching corresponding WBC sample were 20.7% (40 out of 193) for LC cfDNA and 39.1% (18 out of 46) for BLN cfDNA, suggesting that a significant portion of cfDNA variants derive from clonal hematopoiesis, especially in BLN plasma (p = 8.89E-03, chi-squared test).

After filtering for variants potentially derived from clonal hematopoiesis, 153 variants remained in 67 (out of 111, 60.36%) cfDNA samples from LC patients (Figure 2B and Supplementary Table 3), with AFs ranging from 0.03% to 6.00% (median was 0.13%, Figure 2A-C and Supplementary Figure 11). The most frequent variants were missense mutations (n = 102, 67%), followed by nonsense mutations (n = 33, 22%). SNVs (n = 137, 90%) were predominantly C>T transitions (n = 95, 69%) (Figure 2C), which was a feature discovered recently in East Asian LC patients [44]. Mutation frequency analysis revealed that *TP53* was the most commonly mutated gene in LC plasma (mutated in 23% of LC cfDNA samples), consistent with TCGA findings [45–47]. Other frequently mutated genes included *EGFR* (8%), *PTPN11* (8%), *APC* (7%), *APOB* (7%), *KMT2C* (5%), and *KMT2D* (5%) (Figure 2A and Supplementary Figure 12), hence identifying a spectrum mostly consistent with previous reports of lung cancer mutation spectrum (Supplementary Figure 13) [45,48–53]. As for LC subtypes, cfDNA samples from LUSC patients were more frequently muated than that from LUAD patients (Supplementary Figure 13 and Supplementary Table 4).

After stringent QC filtering as well as filtering for WBC-matched variants, as many as 28 mutations remained in 23 (out of 78) BLN plasma cfDNA samples (Figure 2D and Supplementary Table 5), although the percentage of positive samples was much less compared to LC plasma (29.49% *vs*. 60.36%, p = 2.87E-05, chi-squared test). These mutations had AFs ranging from 0.05% to 1.91% (Figure 2A). The median AF (0.13%) was the same as that of LC plasma cfDNA, yet the highest AF was much less (1.91% *vs*. 6.00%). The most frequently mutated genes in BLN plasma were *KRAS* (5%), *CSMD3* (4%), *APC* (3%), *ATM* (3%), *KMT2D* (3%), and *TP53* (3%) (Figure 2D). Notably, 39.3% (11 out of 28) of these were COSMIC hotspot mutations (such as variants affecting the KRAS G12 residue) (Supplementary Table 5). These results suggest that BLN cfDNA harbored common cancer driver mutations, a phenomenon consistent with previous and recent reports that cancer driver mutations are prevalent among normal tissues [43,54,55]. The mutation spectrum of BLN plasma cfDNA is notably different from that of LC plasma: the most frequent mutations found in BLN plasma cfDNA were *KRAS* (5%) and *CSMD3* (4%). *EGFR* variants were never detected in BLN plasma. *KRAS* variants, however, had almost identical frequency in both groups. *TP53* mutations were also observed in BLN plasma cfDNA albeit at a much lower frequency (3%).

### A predictive model based on somatic mutations to distinguish LC from BLN

Next, we asked whether it would be possible to differentiate LC and BLN plasma based on their different cfDNA mutation signatures and AF distribution. To quantify the cfDNA mutational burden, we constructed a mutation score for each cfDNA sample as either a simple summation of the allele fractions of all variants identified therein (SUMAF) or a weighted sum of the allele fractions based on a set of pre-defined weights (weighted SUMAF, or wSUMAF), weighing more on TCGA hotspot cancer driver mutations and COSMIC hotspot mutations, and less on other variants (see Methods for details). We found both scoring methods produced modest classification accuracy for distinguishing LC from BLN plasma: the wSUMAF model generated an area under curve (AUC) value of 0.68, with a sensitivity of 59.5% and a specificity of 71.8% (Figure 2E and Supplementary Figure 14) and the SUMAF model had a similar performance.

### Classification of LC and BLN plasma based on cfDNA methylation data

To identify lung cancer-specific epigenetic changes, such as abnormalities in 5-mC methylome, we performed whole-genome bisulfite sequencing (WGBS) on 25 pairs of LC tissue and normal tissue adjacent to the tumor (NAT) among which 21 pairs were from LUAD, 3 from LUSC and 1 from LCSC (Figure 3A). 315 differentially methylated regions (DMRs) were identified using a cutoff of delta β value greater than 0.2 and p-value less than 0.001 (see Methods), including 293 hyper DMRs and 22 hypo DMRs (Figure 3B). There were a lot more hyper DMRs than hypo DMRs, consistent with the belief that genomic regulatory regions such as promoters of potential tumor suppressor genes undergo remarkable hypermethylation in tumorigenesis. Gene ontology (GO) annotations revealed that the 293 hyper DMRs were significantly enriched for genes encoding DNA-binding domains and homeobox domains, as well as genes involved in the developmental and transcriptional regulation process (Figure 3C), consistent with the possibility that these genes may be involved in lung cancer development by regulating cell differentiation, and when silenced by promoter methylation, may cause cell transformation.

Unsupervised hierarchical clustering using the regional methylation ratio of the identified DMRs perfectly separated LC tissues and NAT with the exception of a single LC sample, highlighting the pronounced epigenetic dysregulation of lung cancer cells. We did not observe notable differences between cancer stages (Figure 3B), consistent with the notion that epigenomic change is an early driver of oncogenesis that persists through later stages of cancer progression.

We next performed comprehensive analysis of 5-mC methylation profile of plasma cfDNA for 111 LC patients and 87 BLN patients using targeted bisulfite sequencing, covering 5.6 million CpG sites located within gene regions, as well as CpG islands, shelves, and shores (Supplementary Table 1). Based on the hyper DMRs we defined using tissue WGBS, we were able to build random forest models that classify LC from BLN plasma (see Methods). To estimate classification accuracy, we performed 10-fold cross-validation (CV), for which AUC was 0.75 (Supplementary Figure 15), a performance slightly better than the mutation-based model. In order to determine whether we could effectively distinguish lung cancer plasma from healthy plasma using fewer DMR markers, we also performed further feature selection. We found that by selecting DMRs with feature importance > 0.008 in each random forest model for each CV, we could achieve a CV AUC of 0.72 (Figure 3D). A total of 76 DMRs were retained in the final CV model. Among cancer patients, the detection sensitivity of the final CV models was higher in LUSC patients than that in LUAD samples (Supplementary Table 6).

### Multi-omics analysis to differentiate LC from BLN plasma

Next, we attempted to integrate multi-omics features to further improve the diagnostic power of our classification model. Indeed, on 91 LC and 71 BLN cfDNA samples that had been sequenced with both targeted deep sequencing and targeted bisulfite sequencing (Supplementary Table 1), we found that combination of methylation features (based on a total of 81 DMRs selected using the same feature selection criteria described earlier) and the SUMAF mutation score achieved an AUC of 0.75, generating a sensitivity of 78.0% and specificity of 60.5% (Figure 4C), compared to an AUC of 0.68 achieved by mutation score alone (Figure 4A), and an AUC of 0.72 achieved by methylation features alone in the same set of samples using the same CV procedures (Figure 4B).

In addition, levels of 5 serum marker, CEA, CYFRA21–1, NSE, CA19–9, and CA125, were also measured in a subset of the blood samples (Supplementary Table 1). We found that among the five protein markers, only CEA level appeared to be significantly higher in LC patients than BLN patients (p = 0.0438), producing a modest AUC of 0.66 for classifying the two groups (Supplementary Figure 16 and 17). Therefore, we tried to incorporate CEA into the predictive model in addition to the DMR features and mutation score in samples with complete measurements (74 LC and 60 BLN samples). The multi-omics predictive models based on SUMAF mutation score, top DMRs (a total of 81 regions) and serum CEA level achieved an AUC of 0.79, with 75.7% sensitivity and 68.3% specificity (Figure 4E), which showed further improvement compared to the model without CEA on this set of samples (AUC = 0.75) (Figure 4D). Similar to classification models based solely on mutation or methylation, higher prediction accuracy was found for LUSC patients than LUAD patients in integrated bi-omics and multiomics models (Supplementary Table 6).

### cfDNA mutational burden and methylation level as prognostic factors for lung cancer

We first tested whether mutational status (wSUMAF, < 0 *vs*. > 0) was associated with lung cancer overall survival (OS) among lung cancer patients. We found that among lung cancer patients, the high mutational burden was associated with a significantly worse OS (Figure 5A). Notably, among stage I lung cancer patients, a significant association was observed between mutation score and OS (Supplementary Figure 18). To further improve the performance of model prediction on lung cancer prognosis, we attempted to identify potential methylation-based prognostic biomarkers and incorporate these features into the model(Supplementary Figure 19). We divided the LC cases into training and testing set and applied a DMR selection procedure on the training set using the penalized COX regression, which identified 12 DMRs that were potentially associated with lung cancer prognosis and obtained corresponding coefficients (Supplementary Table 7, see Methods for details on the analysis procedure). Of these, DMRs of gene *FOXG1*/*LINC01551, TMEM240, AKR7L, CBLN4*, and *GCK*/*MYL7* appeared to be associated with a worse lung cancer prognosis while DMRs of *PRDM11, LOC440028/SBF2-AS1, GFI1*, and *ST3GAL1*were associated with a better prognosis. We then calculated a methylation-based prognostic score (MPS) for each individual as the sum of the products of the DMR methylation level and corresponding coefficient. We thereafter experimented with combining the mutation score with the MPS as the bi-omics prognosis score and tested its association with survival. Patients with a high mutational burden and a high MPS were categorized as the high prognosis score group, while other patients were categorized as the low prognosis score group. Compared to patients with a low prognosis score, patients with a high prognosis score had a significantly worse OS in the testing set (Figure 5B). Finally, to avoid information loss due to categorization, we modeled both mutation score and MPS continuously (see Methods for details) and built two separate multivariate Cox proportional hazard models on wSUMAF only, as well as on combined wSUMAF and MPS with adjustment of age, stage, histological type and smoking status. Higher AUCs were obtained using the bi-omics prognosis model than the mutation only model (Supplementary Figure 20). Taken together, these results suggest that integrated genomic features have the potential to be used as better prognostic markers for lung cancer.

**Figure 5:**
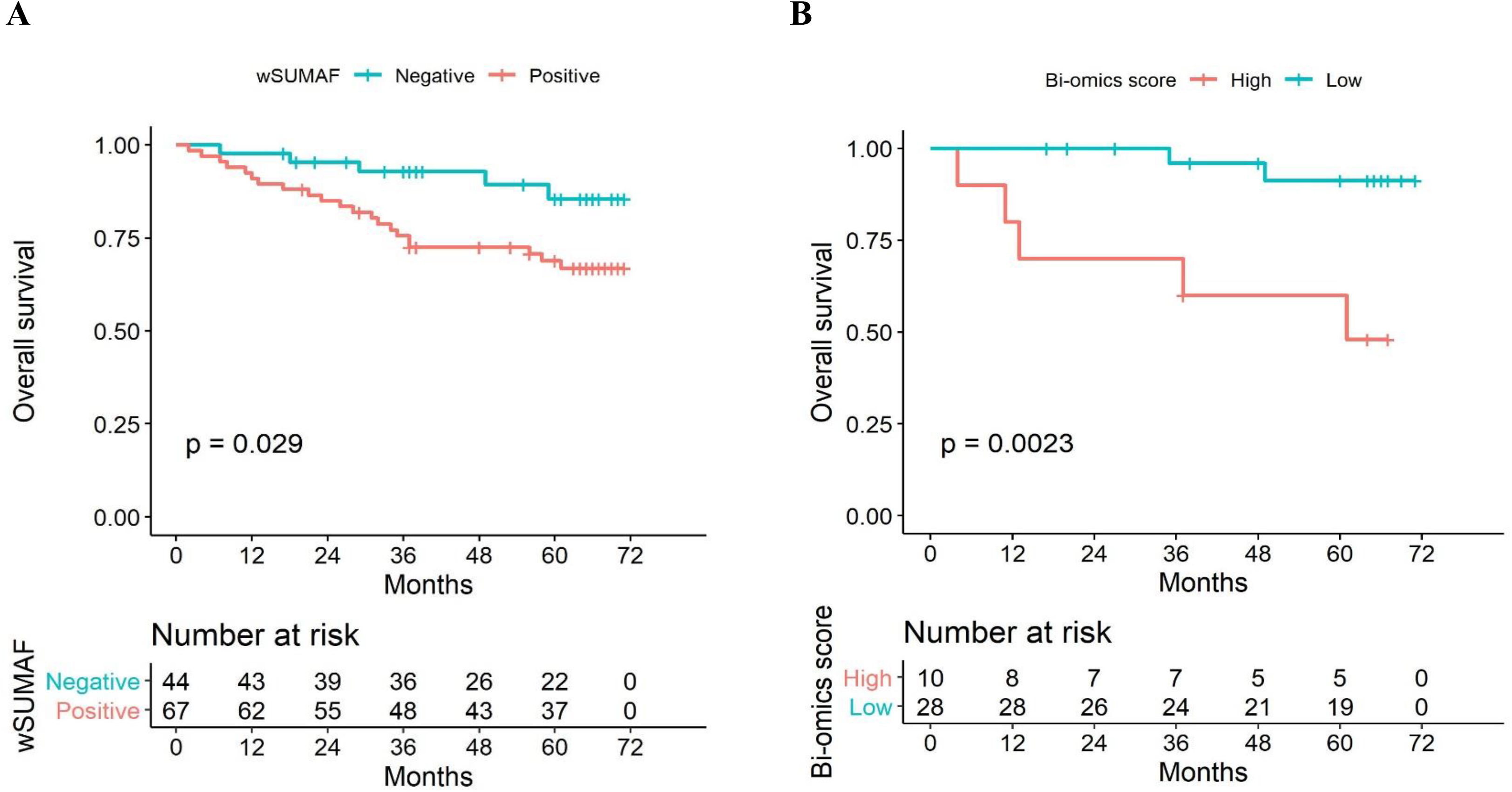
Kaplan-Meier plot on omics-based lung cancer prognostic model in relation to OS. (A) Mutation scores in the whole dataset; (B) Multi-omics scores in the testing set.

## Discussion

In this study, we applied targeted ultra-deep sequencing to plasma cfDNA of LC and BLN patients. Matched WBC DNA were sequenced in parallel, which revealed that non-synonymous variants were prevalent in WBC DNA for both the LC and BLN group. Further analyses showed that a notable portion of cfDNA variants was detected in matched WBC (20.7% for LC plasma, and 39.1% for BLN plasma) and their VAFs were well correlated (Figure 1E), suggesting that these variants most likely derived from WBCs. VAFs of the majority of WBC-matched somatic mutations detected in the cfDNA were less than 1%, hence would have been missed if WBC DNA has not been sequenced with ultra-deep sequencing. These results corroborate recent findings that WBCs, carrying variants accumulated through clonal hematopoiesis (CH), constitute an important source of somatic mutations found in cfDNA [43]. Notably, some of the shared variants between cfDNA and matched WBC samples were cancer hotspot mutations (Supplementary Figure 10), suggesting that CH variants may indeed significantly confound cfDNA analysis if not analyzed in parallel. Interestingly, *TP53* variants were never detected as shared variants between BLN plasma cfDNA and matched BLN WBC sample (Supplementary Figure 9). This may indicate that the accumulation of TP53 mutations through CH may be somehow related to the cancer risk of the individual.

After removing WBC derived variants, we found that cfDNA mutations were prevalent in BLN plasma cfDNA (29.5% of samples analyzed contained at least one variant). This finding is consistent with recent studies showed that benign tumors may also harbor somatic mutations, including those in cancer driver genes [56,57]. However, because we did not obtain matched BLN tissue for these plasma samples, it remained to be determined whether the mutations found in BLN cfDNA could be attributed to mutations that may have arisen in the benign lesions of the lung. If this indeed is the case, then we would expect it to be challenging to classify malignant and benign disease solely based on cfDNA mutational status. Further study would be needed to clarify whether patients bearing benign lung lesions indeed have a higher mutational burden in their cfDNA than healthy individuals.

Not surprisingly, predictive models built on mutation score alone had limited classification ability for distinguishing between LC and BLN plasma (AUC = 0.68). Some earlier studies suggested that mutational status can be used to diagnose LC from benign lung nodules with high specificity and modest sensitivity [58,59], but these conclusions may have suffered from potential bias caused by limited sample sizes used in the study. Our results were obtained from a larger sample size (128 LC and 94 BLN plasma) and showed that diagnostic model based on mutational status alone had sub-optimal classification accuracy and hints that a multi-analyte approach is more likely to improve the detection of cancer signal.

By performing WGBS on lung cancer tissues and NATs, we identified more than 300 lung cancer-specific DMRs, with the majority of them being hypermethylated DMRs, suggesting that hypermethylation of genome regulatory regions is an important event in lung cancer development. Indeed, these DMRs are enriched for genes involved in transcriptional regulation and are likely to cause profound downstream changes in gene expression and contribute to cell transformation; these genes are likely to be potential tumor suppressor genes, and many of which haven’t been implicated as such previously (such as *SEC31B, ZNF274*, and *NXPH1*). A small number of DMRs are hypomethylated, and therefore may encode potential cancer driver genes. To our knowledge, this is the first time many of these genes are implicated in epigenetic dysregulation of lung cancer. DAVID functional GO analysis of biological processes revealed that these DMR genes were enriched in skeletal system/embryonic organ development/morphogenesis (Supplementary Figure 21), indicating that the lung cancer cells may have obtained some characteristics of embryonic stem cells. Functions of these genes remain to be elucidated in further studies and may help us better understand the underlying molecular mechanisms of lung cancer development and progression.

Our cfDNA methylation-based classification model for LC and BLN plasma achieved a slightly better cross-validation AUC (0.72) than the mutation-based model, suggesting that LC-specific methylation changes are potentially useful markers for diagnosing lung cancers versus benign lesions. Further multi-center studies with larger sample sizes will be needed to validate the utility of selected markers and the robustness of our diagnostic model. We also noted that performance of our models are slightly inferior to an earlier study which used a panel of 9 methylation markers for differentiating early-stage lung cancers from benign pulmonary nodules (AUC of 0.82 (0.70–0.93) in the test set), even though we used more methylation markers in our model. The difference in model performance could be attributed to the different study populations and cfDNA analysis methods. The smaller cohort size in the previous study may have also caused over-fitting and/or over-estimation of the model performance. The discrepancy also suggests that we need to be cautious with the development and validation of such cfDNA-based diagnostic models, considering the intrinsic technical difficulties of detecting a minute amount of cancer-derived signals in circulation and relying on machine-learning approaches to build diagnostic model, a process that can be heavily affected by batch effect as well as variations in sample characteristics, especially with single-center clinical study.

In our results, we found that a higher percentage of LUSC cfDNA samples were mutation positive than LUAD samples (Supplementary Table 4). We also observed higher sensitivity for detecting LUSCs than LUADs using methylation as well as multi-omics based classification models (Supplementary Table 6). These results are consistent with the notion that LUSCs are significantly more necrotic than LUADs and are more likely to shed ctDNA into circulation [60].

Previous studies have shown that detection of cancer driver mutations in cfDNA, when combined with serum protein markers, can be used to increase sensitivity without significantly sacrificing specificity for cancer detection [9,61]. In our study, the multi-omics model integrating mutation, methylation and serum protein marker further improved the performance of the classification model (AUC of 0.79), compared to the mutation-based model or methylation-based model. To our knowledge, this is the first proof-of-concept study to demonstrate that genetic, epigenetic, and proteomic analytes could be combined to increase the performance of liquid biopsy-based diagnostic model for lung cancer. Further study with a larger size of clinical samples will be needed to validate the robustness of this approach.

We also investigated the association of prognosis of lung cancer patients with cfDNA mutation and methylation status. Lung cancer patients with any mutation were observed to have an unfavorable outcome compared with those without, in line with previous studies [62–64]. We also identified a group of potential methylation-based lung cancer prognostic markers from the pool of lung cancer tissue-specific DMRs and constructed a methylation-based prognostic score (MPS). Previously, multiple methylation-based prognostic classifiers had been reported for lung cancer, however, the reported markers were mostly inconsistent [25,65–67]. The inconsistency could be explained by limited sample sizes, variations in study design, as well as different detection methods used. Further studies will be needed to validate the clinical utility of these markers including ones discovered in the current study. Finally, we found that combining continuous MPS with the mutation score could improve the prognostication model compared with the multivariate model based solely on mutation. One potential caveat to be noted here is that since the estimated coefficients of DMR markers for generating the MPS might not be accurate enough due to relatively limited sample size; therefore, additional study with larger sample size would be necessary to validate current findings. Overall, we provided proof-of-principle evidence that combination of multiple blood-derived biomarkers has the potential to improve lung cancer prognostication.

## Availability of Data

The data reported in this study are alsoavailable in the CNGB Nucleotide Sequence Archive (CNSA: https://db.cngb.org/cnsa; accession number CNP 0001236).

## Acknowledgements

This study was supported by the National Natural Science Foundation of China (No.81602001), Peking University People’s Hospital Research and Development Funds (RS2019–01), and Shenzhen Engineering Laboratory for Innovative Molecular Diagnostics (DRC-SZ[2016]884).

